# Dynamic Mortality Risk Prediction in Myelodysplastic Syndromes Using Longitudinal Clinical Data

**DOI:** 10.1101/2025.07.21.25331775

**Authors:** Jonathan Bobak, Philipp Spohr, Sarah Richter, Alexander Streuer, Felicitas Schulz, Corinna Strupp, Catharina Gerhards, Nanni Schmitt, Thomas Luft, Sascha Dietrich, Ulrich Germing, Gunnar W. Klau

## Abstract

**Purpose:** Patients with myelodysplastic syndromes (MDS) exhibit diverse disease trajectories necessitating different clinical approaches ranging from watch-and-wait strategies to hematopoietic stem cell transplantation. Existing risk scores like the IPSS-R or EASIX provide static risk stratification at diagnosis but do not capture evolving disease dynamics. We address this problem by introducing a dynamic, data-driven approach to predict short-term mortality risks repeatedly, across the patient’s disease course.

**Materials and methods:** We develop a machine learning model based on gradient-boosted decision trees to estimate one-year mortality risks from both longitudinal parameters from blood values and diagnosis-based features. We train the model on a dataset of patients from the MDS Registry Düsseldorf (n=1024) and validate on patients from University Hospitals Heidelberg (n=286) and Mannheim (n=31).

**Results:** Validations on independent cohorts achieved AUROC scores of around 0.8 and better predictive performance for one-year survival compared to a diagnosis-only baseline model. The model accurately predicted mortality risks as early as within the first 90 days of diagnosis. Feature importance analysis revealed clinically plausible feature-label relations, supporting interpretability.

**Conclusion:** This dynamic risk model enables continuous, individualized assessment of one-year mortality risk in MDS patients, offering a supplement to static scores used at diagnosis. Our results highlight the utility and importance of including longitudinal parameters in risk assessment analysis.

## Introduction

Myelodysplastic syndromes (MDS) are a class of malignant stem cell diseases resulting in reduced and defective blood cells. The main affected age group are elderly patients, with survival times ranging from a few months to multiple years [1]. Quantitative mortality risk assessment for this diverse set of clinical trajectories is paramount in clinical decision-making. The main approach for MDS in recent years has been to focus on diagnosis-based modelling. Scores like the International Prognostic Scoring System are broadly applied in the form of the revised and molecular IPSS-R/IPSS-M [2], [3] as well as other scores calculated on diagnosis [4], [5] relying on the same methodology. A major advantage is the simplicity of calculating them. For the IPSS-R at diagnosis just five parameters are needed to classify a patient into one of five categories ranging from very low to very high risk. But this simplicity and focus on a single point in time leads to some drawbacks. Losses in predictive power over time due to similar hazards in all risk categories were observed [6]. Additionally, the categorization effectively limits predictions to five hazard profiles. For an individual patient, the question if they do follow their corresponding risk profile at any given time cannot be answered.

For longitudinal predictions, clinicians must rely on their experience since re-evaluation of the diagnosis-based scores is not always possible and may require more intensive diagnostics rarely performed on follow-up. The WHO-based prognostic scoring system (WPSS) [7] investigated a time-adjusted score with good predictive performance for overall survival and AML progression but is still limited to 5 risk categories. Longitudinal data also comes with lots of challenges like unevenly sampled or missing data and more complex patterns. To approach these challenges prediction tools based on machine learning algorithms are a major focus to recognize patterns and improve predictions. Orgueira et al. applied random survival forests to improve model performance compared to the IPSS-R [8]. This study did not only focus on individual predictions instead of general risk classifications but also showed the advantages of complex, non-linear models to learn new patterns. Tree boosting was also used to predict MDS diagnosis risks one year prior to clinical diagnosis with good classification performance [9]. Apart from focusing on a single point in time as a basis for model prediction, dynamic predictions have been applied to analyse 100-day mortality risks after hematopoietic stem cell transplantation. Risks are predicted each day starting on the day of transplantation up until day 30 showing that with increasing longitudinal data predictions improved [10].

Here we present an approach with a moving one-year prediction horizon in relation to the time of prediction. This conceptual approach for continuous and event independent predictions and its caveats were investigated by Sherman et al. [11]. Our model leverages both diagnosis-based and longitudinal features as input. We trained and validated the model on retrospective data from the MDS Registry Düsseldorf and retrospective validation datasets from the University Hospitals Heidelberg and Mannheim showing promising prediction improvements by leveraging longitudinal data in addition to diagnosis-based parameters. Our model may serve as a valuable supplement to existing risk scores, especially for longer follow-ups and dynamic disease trajectories. A conceptual overview of our approach can be seen in Figure 1.

**Figure 1:**
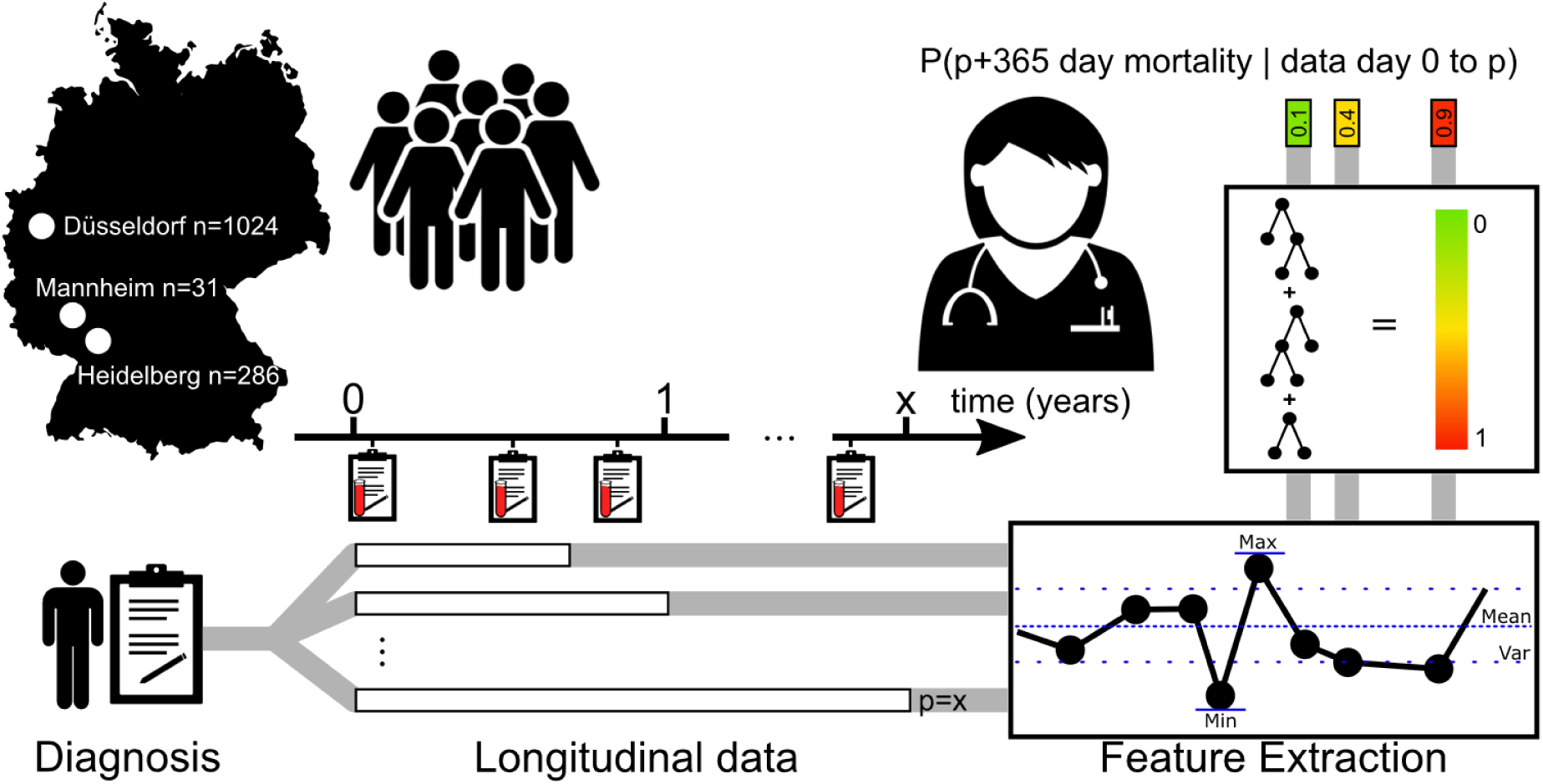
Overview of our study pipeline. We use the Düsseldorf patient dataset for training and initial validation of our method supplemented by two validation datasets from the University Hospitals Heidelberg and Mannheim. For each patient, we gather both diagnosis-based and longitudinal features. For the retrospective longitudinal data, we sub-sample each patient at different time points and extract a standardized set of features. Combined with diagnosis-based features these serve as input for a gradient-boosting model which predicts a probability for one-year mortality for each sub-sample. A higher predicted probability is interpreted as a higher risk of dying within 365 days after the time of prediction.

## Materials and Methods

### Patient inclusion criteria

Our inclusion criteria for patients, related to technically necessary minimal data availability and bias reduction, are

- surviving at least six months
- receiving no hematopoietic stem cell transplantation (HSCT) at any time, as this procedure is altering the natural course of the disease
- having at least three values for each longitudinal variable
- having at least one year of follow-up from each prediction point in case of right-censored patients

By setting a minimal survival time for inclusion we aim to alleviate the leading bias for very high-risk patients being diagnosed very late in disease progression. This does not fully eliminate the bias but allows us to focus on more subtle, long-term disease patterns.

### Conceptual framework

We follow the framework described by Sherman et al. [11], defining a fixed reference point *p*_0_ as the time of diagnosis. Each prediction *p*_*i*_at time *i* incorporates all available data between *p*_0_and *p*_*i*_, estimating the probability of death within 365 days from *i*.

This setup ensures that predictions are temporally localized, incorporate full clinical history up to time *i*, and are independent of prior predictions and the outcome itself. While prediction windows may overlap, each instance is treated independently, which is crucial for downstream performance evaluation. Successive predictions advance the prediction horizon.

### Data pre-processing and feature extraction

Patient data has two modalities. Diagnosis-based features are only observed at *p*_0_while longitudinal time-series features are measured repeatedly. The longitudinal information is irregular and sparse as patients were monitored in a real-life clinical environment. Each patient’s complete retrospective longitudinal data is subsampled each quarter with observations. Quarterly sampling was chosen such that densely observed hospitalization periods are not overrepresented, but a representative set of intermediate prediction points is obtained. The point *p*_*i*_ is the last observation within a quarter. Due to decreasing numbers of data points, sampling was stopped after 32 quarters.

To prevent label leakage, we exclude the final 60 days before death from sampling, as this period may overshadow more subtle early signals of deterioration during training. From the irregular and sparse longitudinal data, we extract a standardized set of characteristic features for each subsample (see Supplementary Table B). These derived time-dependent features are then combined with the fixed diagnosis-based features to construct the final input vector for each prediction instance.

Samples are labelled positive (1) if death occurs 365 days after *p*_*i*_or negative (0) otherwise. By design, a patient may have samples with different labels which are obtained independently of one another as they represent different prediction times but may share similarities in features.

### Model Architecture

Our longitudinal survival prediction model using XGBoost [12] incorporates both diagnosis-based and longitudinal features. The input features used for model training are summarized in Table 1. The longitudinal features were mainly chosen due to their frequent observation in the clinical setting at both ordinary follow-ups and adverse events. Similar, bone-marrow blasts and cytogenetics were only included at diagnosis as follow-ups to these variables are biased to clinical deterioration and disease progression. The IPSS-R [2] was specifically excluded from the feature set since most of its constituent variables are already model inputs. We also do not consider therapies as inputs due to uncertainty around dates and a huge variety of treatment schemes. The model has to learn related patterns implicitly and may cope better with new combinations of drugs or new drugs since the input features do not depend on them. Mutational data could not be incorporated due sparse and incomplete data.

**Table 1:**
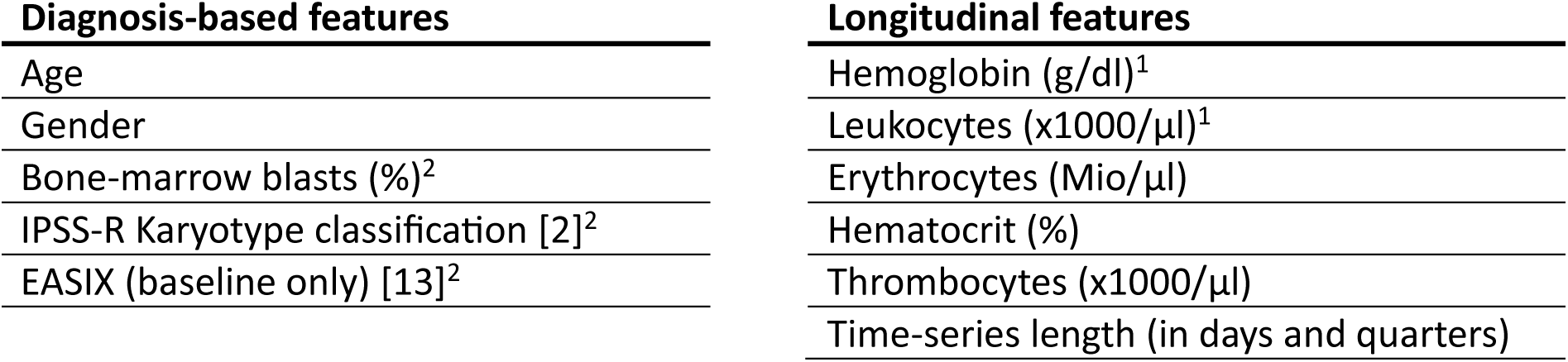
Input features for model training. Diagnosis-based parameters have singular values while the longitudinal features correspond to series of observations. EASIX is just used for the baseline model. The survival time is input for both models and is given in fine-grained days as well as 90-day quarters. ^1^ Both Hemoglobin and Leukocytes are present in the baseline and longitudinal model. For the baseline model the measurements at diagnosis, for the longitudinal model a series of measurements is used. ^2^ These features are optional, i.e. the models can handle missing values for them, at least one feature is required.

| Diagnosis-based features | Longitudinal features |
| --- | --- |
| Age | Hemoglobin (g/dl)<sup>1</sup> |
| Gender | Leukocytes (x1000/μl)<sup>1</sup> |
| Bone-marrow blasts (%)<sup>2</sup> | Erythrocytes (Mio/μl) |
| IPSS-R Karyotype classification [2]<sup>2</sup> | Hematocrit (%) |
| EASIX (baseline only) [13]<sup>2</sup> | Thrombocytes (x1000/μl) |
|  | Time-series length (in days and quarters) |

As a comparative baseline, we also train a model only on diagnosis-based features. Contrary to the longitudinal model, we included the EASIX (Endothelial Activation and Stress Index) score [13] as a replacement for raw thrombocyte counts.

The XGBoost implementation handles missing data natively by adding additional decision branches, when a value is missing, which allows us to retain patients with missing diagnosis-based features. Longitudinal features are always present. Hyperparameters are selected via grid search with cross-validation on the training set. To reduce overfitting given the modest dataset size, we apply regularization strategies and minimize false negatives by tuning model sensitivity with sample weights during training (see Supplementary Table C).

### Evaluation Methodology

Classification performance is evaluated using three metrics: area under the receiver operating characteristic curve (AUROC [14]), area under the precision-recall curve (AUPRC[15]), and Brier Score [16]. The AUPRC is particularly relevant due to the observed class imbalance, which the AUROC metric has been shown to omit [15], [17]. The Brier Score quantifies absolute error in relation to the true probabilities.

For a robust initial model performance assessment, we conducted repeated cross-validation on the Düsseldorf training cohort. Metrics were averaged over all cross-validation folds to account for variability due to cohort composition and to assess general model capability. Changes in performance for predictions further from diagnosis were evaluated by calculating metrics within each quarterly bin.

Model performance was compared to the established IPSS-R [2] using the same cross validation strategy. For a fair comparison we trained and evaluated on a sub-cohort of patients for which the IPSS-R was present. Using the IPSS-R category and conditional Kaplan-Meier curves [18], [19] of the training sets we obtained longitudinal predictions for each sample. A comparison with the more recent IPSS-M [3] was not possible since too little genomic information was available.

To evaluate generalizability across institutions and real-world applicability, we trained our model on the Düsseldorf dataset and evaluated it on two independent validation cohorts from University Hospitals Heidelberg and Mannheim.

Finally, we performed feature importance analysis on this generalizable model using the average reduction in impurity achieved for each feature over all trees and SHAP values [20] a game theoretic approach to explain relations between input features and model outputs.

### Datasets

The training cohort consists of 1024 retrospective MDS patients treated at the University Hospital Düsseldorf, all part of the Düsseldorf MDS Registry, with a total of 6146 samples and a 2.66:1 ratio between negative to positive labels. We validated on two retrospective cohorts from the University Hospital Heidelberg (patients=286, samples=1708, label balance ≈4:1) and the University Hospital Mannheim (patients=31, samples=237, label balance ≈10:1). For the validation sets, we did not filter the final 60 days of longitudinal data to simulate real predictions without event knowledge.

Sample numbers diminish over time with similar decrease across datasets and labels (see Supplementary Figure A). The Heidelberg cohort shows a lower median follow-up and higher mortality than the other datasets indicating more high-risk patients with earlier events but similar average sample numbers per patient. Detailed demographics and characteristics can be found in Supplementary Table A.

### Software Availability

The pipeline used to train and evaluate the model is written in python using snakemake [21] as a workflow library for easy reproduction. Additionally, we used tsflex [22] and tsfresh [23] for feature extraction, XGBoost [12], SHAP [20] and scikit-learn [24] for model training and evaluation, Pandas [25], [26] and NumPy [27] to handle and transform data, and matplotlib [28] for plotting. All code is available here: https://github.com/dietrichlab-cs/dynamic_one-year_mortality_mds (archive: <u>DOI: 10.5281/zenodo.16102058</u>). A small webtool, to test both models, is available (https://dietrichlab.de/PythonApps/dynamic_mds_paper/).

## Results & Discussion

### Adding longitudinal data improves predictions

We first trained the baseline and longitudinal model on the Düsseldorf cohort and evaluated our metrics on the external validation cohorts from Heidelberg and Mannheim. Table 2 shows results for all metrics and datasets comparing models. On the Heidelberg dataset, the longitudinal model improves the AUROC by approximately 0.12 and the AUPRC by 0.23. Both models perform substantially better than random guessing, which would yield an AUROC of 0.5 and an expected AUPRC of around 0.2. Furthermore, the longitudinal model exhibits a lower Brier score than the baseline, indicating improved overall calibration and probabilistic accuracy.

**Table 2:** AUROC, AUPRC and Brier Score for all three datasets. For the Düsseldorf cohort averages from cross validation are given. For the Heidelberg and Mannheim datasets performance is measured on models trained on all patients from Düsseldorf. Results are rounded to four decimal places.

| Dataset | Metric | Baseline | Longitudinal |
| --- | --- | --- | --- |
| Heidelberg cohort | AUROC | 0.7232 | **0.8364** |
|  | AUPRC | 0.4873 | **0.7057** |
|  | Brier Score | 0.2035 | **0.1519** |
| Mannheim cohort | AUROC | 0.6391 | **0.8552** |
|  | AUPRC | 0.2757 | **0.5820** |
|  | Brier Score | 0.2022 | **0.1986** |
| Düsseldorf cohort<br>(cross-validation<br>mean) | AUROC | 0.6850 | **0.8103** |
|  | AUPRC | 0.4600 | **0.6187** |
|  | Brier Score | 0.2147 | **0.1691** |

For the Mannheim cohort similar improvements for the longitudinal model can be seen for the AUROC and AUPRC scores with improvements of 0.22 and 0.31. Interestingly, Brier scores for this dataset do not show the same difference but are close across models yet in the same range as for the Heidelberg and Düsseldorf datasets.

The baseline model does perform equally good as the IPSS-R with the advantage of being able to cope with missing values shown in Supplementary Figure C. The longitudinal model outperforms both other models clearly. All subsequent comparisons are done between the baseline and longitudinal model as they allow for larger patient cohorts.

The intuitive assumption, that adding information from longitudinal data does improve model performance compared to an approach solely reliant on data from the time of diagnosis, could be confirmed. We can conclude that our longitudinal model trained on patients from Düsseldorf does indeed learn generalizable patterns and is applicable to other, real-world cohorts.

### Longitudinal model can react to dynamic changes

We do see differences in the two models approach to predicting risks. The baseline model is restricted to an initial state of a patient and their survival time. An initial prediction based on the diagnosis features is made which is then adjusted according to survival time. Thus, a major factor is the correlation between survival time and mortality. This approach is very similar to classical survival analysis in the form of Cox-regression or Kaplan-Meier curves [19], [18]. Predicted risks of different subsamples in one patient generally monotonically decrease over time. Figure 2 shows some example patients and their predictions for both models as well as the ground truth labels. Patients A and B start with initial risks gradually decreasing. For patient A this is correct, as we observe no positive sample. While patient B does die shortly after the 21st quarter but the baseline model predicts a low risk. For patient D the predictions indicate low risk from the start although the patient only has positive samples. The longitudinal model does predict dynamically as it does receive new and changing longitudinal data. For patient B we see an increasing risk over time with the highest probabilities on the last two predictions 75 and 3 days prior to the patient dying. The same observation can be made for patients C and D including a higher initial risk for patient D compared to the baseline model.

**Figure 2:**
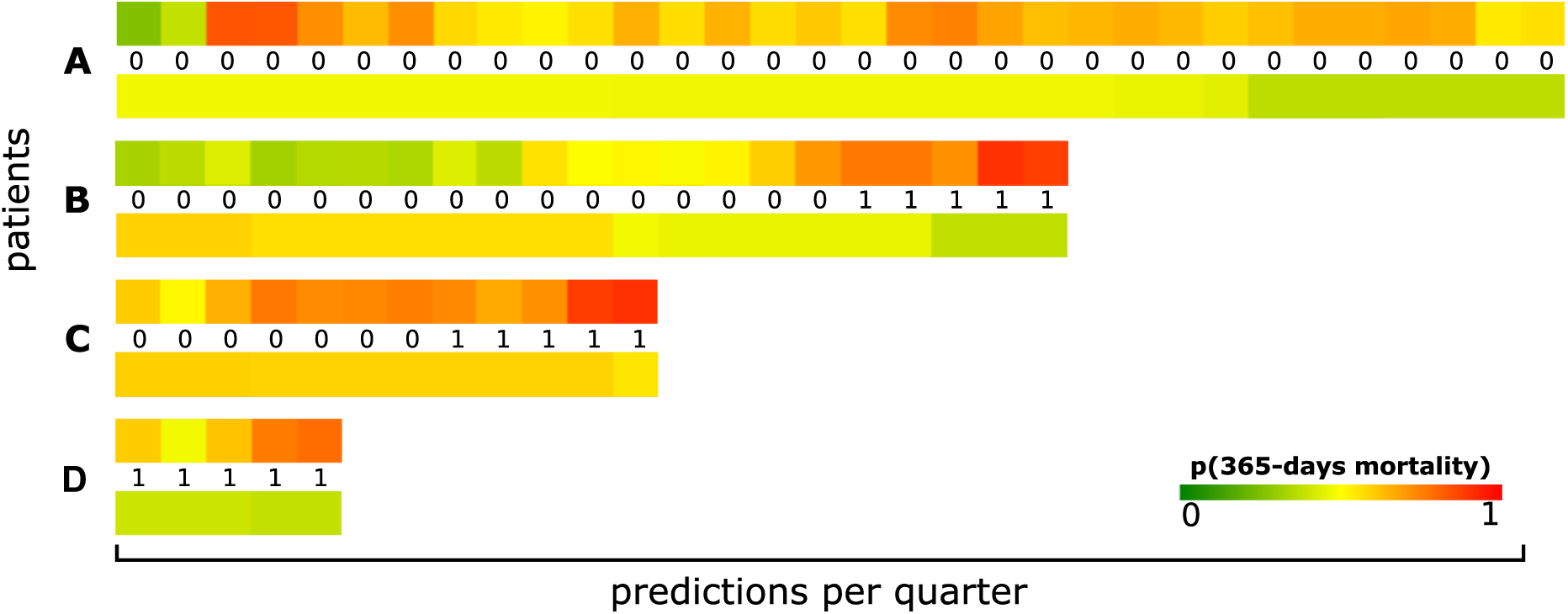
Example predictions for four different patients. The numbers between the coloured bars correspond to the true label for each prediction, the top bar displays the probabilities given by the longitudinal model, the bottom bar those given by the baseline model. Green is associated with likely survival while red indicates a high risk for dying 365 days after the time of prediction.

Dynamic predictions due to added information from longitudinal features are advantageous while the baseline model is bound to survival time alone and is subject to population dynamics resulting in probabilities being independent of a patient’s actual disease trajectory similar to classical survival curves.

### The longitudinal model learns generalizable patterns

For the two external validation sets we explored how the longitudinal model predicts samples within the last 60 days before the occurrence of a mortality event. These samples would not have been included in training. For both datasets and all samples falling under this condition, the longitudinal model predicted either higher or equal probabilities for mortality compared to the prior prediction. Examples are patients B and C in Figure 2. Both patients die within the last quarter of predictions thus the last two predictions are of interest. They have the highest predicted risk, clearly indicating imminent mortality. We can see that the patterns of longitudinal features right before the mortality event are either similar, more pronounced or continuations of patterns from earlier prediction points.

We investigated other endpoints in MDS, namely AML progression and transfusion burden. For AML progression, the model did report higher mortality risks around the time of recorded progression for most patients in both validation cohorts. This time point is the clinical validation of progression, making it hard to assess how well the model serves as an early warning. A similar uncertainty arises when looking at transfusion burden. Patient A in Figure 2 received transfusions over the whole observation period. As a side-effect a study inclusion, investigating iron overload, was checked. This may explain why the longitudinal model predicts a constant risk for this patient as 8 years of close clinical monitoring and regular transfusions do not hint at a low-risk trajectory. Although the model seems to associate higher risk with specific events it is not conclusive if the model can act as an early warning or if it just reports on side effects of more aggressive therapies/disease progression.

### Trade-off between false positives versus false negatives

As seen exemplary for patient A in Figure 2 the longitudinal model is prone to false positive predictions. One reason for those may be medical aspects relating to therapy and disease progression. Another influencing factor was our deliberate choice to introduce sample weights during training increasing the penalty for false negative predictions, pushing the model towards a high recall for positive samples, evident by the achieved AUROC scores.

Over both validation datasets we qualitatively investigated false negatives. Many of these could be attributed to either short-term mortality or events unrelated to MDS. Some examples include septic shock after hip surgery or liver failure due to liver cancer. Although MDS is a contributing comorbidity, if the resulting cause of death is not directly visible within the longitudinal data, our model cannot predict these events. This may occur if the patient was treated elsewhere, the blood values were not available for evaluation, or the cause of death does not affect blood values as evident in the training set.

### Model performance does not depend on sample length

To investigate predictive power over time, we perform cross-validation on the Düsseldorf dataset, tracking evaluation metrics for each split and sample length and average them. All metrics show a similar trend (see Figure 3). The difference between the longitudinal and baseline model starts out very small and then increases over the first quarters up to roughly the 12-13th quarter with the longitudinal model outperforming the baseline. Overall, the longitudinal model stabilizes around an AUROC of 0.8 and an AUPRC of 0.6 with improving Brier scores but is getting more unpredictable for longer sampling periods. The baseline model shows a decreasing performance trend for both AUROC and AUPRC with a constant Brier score apart from the last 4-5 quarters which do show an overall increase in model performance.

**Figure 3:**
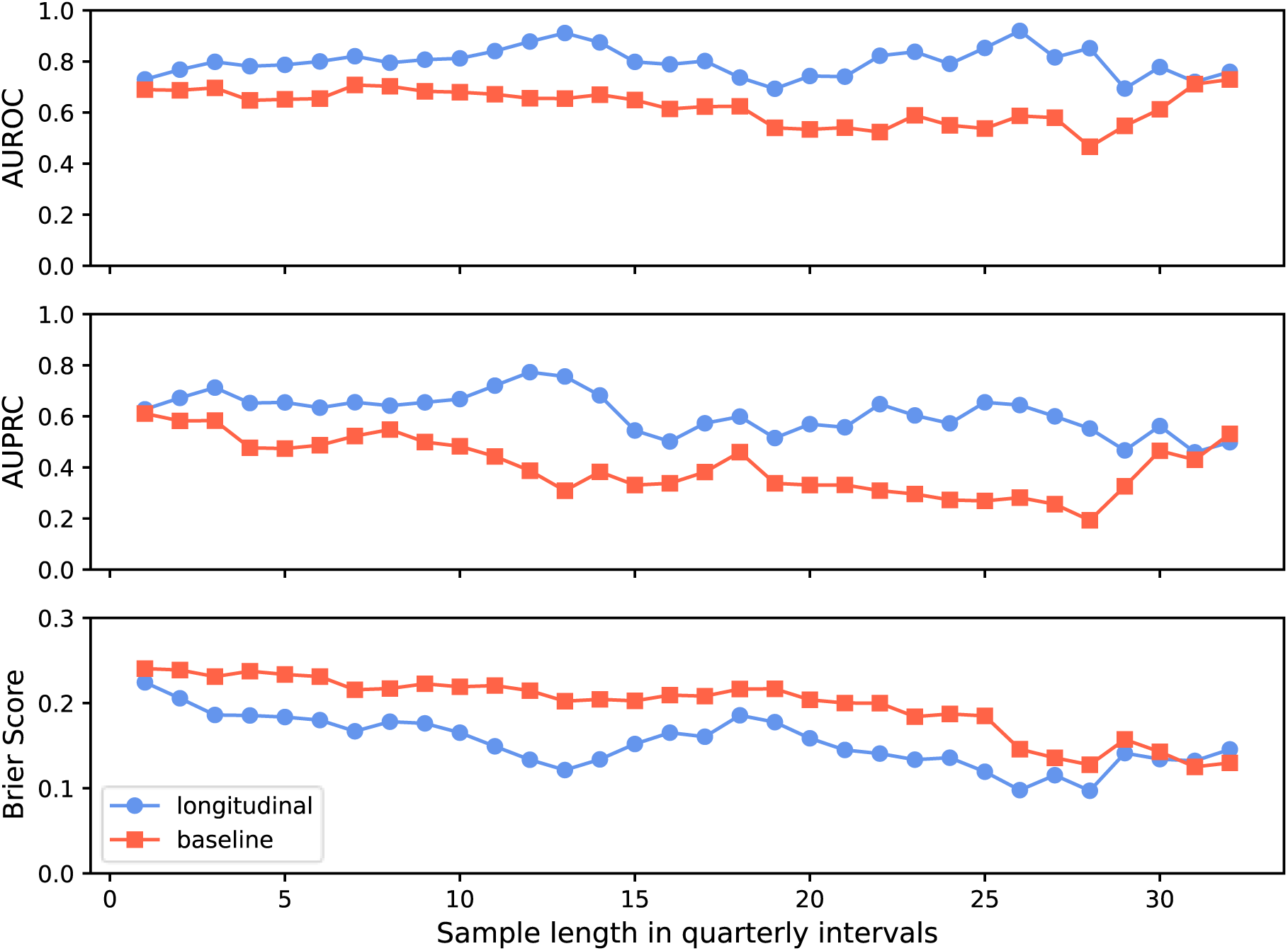
Average evaluation metric values when performing cross-validation on the Düsseldorf dataset. For AUROC and AUPRC higher, while for the Brier Score, lower values are better. See Supplementary Figure B for information on variance across cross-validation runs. Increasing variance and reduced sample size are reasons for both models converging to similar performance. See Supplementary Figure B for more details on result variance.

As both models start out on similar data even without lots of longitudinal data, the longitudinal model performs a lot better. We cannot completely rule out dataset specific variances and biases as reasons for some of the performance trends but the fact that the differences in model performance are persistent and we do observe independent increases and decreases between models across different cross-validation folds leads to the conclusion that the longitudinal model learns generalizable trends. One caveat for this analysis is the high variance across splits (see Supplementary Figure B) due to diminishing amounts of samples and changes in label balance. However, this variance is consistent across both models supporting observations on average performance.

The early advantage of the longitudinal model indicates that disease dynamics hold predictive value early on, stressing the importance of continued quantitative monitoring of risks.

Meaningful feature – label relations

Feature importance evaluations show meaningful feature-label relations validated by medical domain experts for the 20 most important features as seen in Figure 4. As an example, lower hemoglobin averages indicate a higher associated probability of mortality. According to the MDS treatment strategy, patients with lower hemoglobin values will need transfusions [29]. Additionally, hemoglobin has been shown to be a robust marker for risk assessment [2], [3]. We can also observe 7 out of 20 most important features relating to quantile distributions across all longitudinal features. This may be indicative of the model learning to detect outliers and assess mortality risk based on this. Adding to this hypothesis is the inclusion of variation coefficients and averages in this list. Patients which are in regular need of transfusion will not only have more data to base the prediction on but also show more variance. Since complete data on exact treatments was not broadly available, we can only speculate that the model does pick up on treatment related patterns like transfusion burden. There has been evidence that increased transfusion-need is correlated with worse outcome [5], [30]. One more feature type we can observe in higher abundance is the slope of the last three data points of various longitudinal features. Even though this is not an outlier, it is a feature type capturing a more localized view of a patient’s recent disease history including short term changes. According to SHAP analysis negative slopes are associated with higher risk.

**Figure 4:**
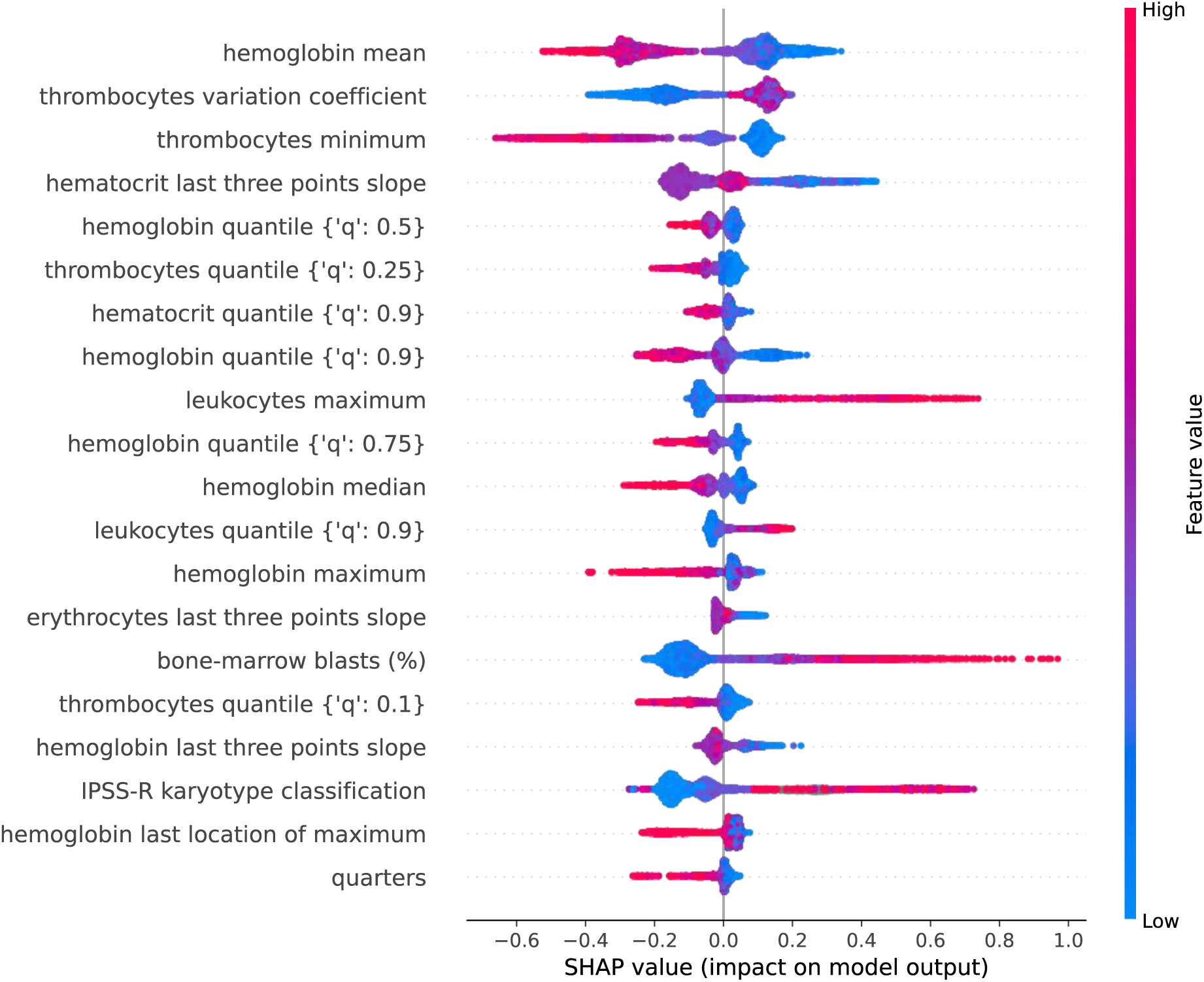
SHAP values for the 20 most important features according to the XGBoost “gain” metric. Gain describes the average reduction in loss when a feature was chosen in a split across all trees and training samples. For each feature and sample, the SHAP value gives an intuition if the feature biases the prediction towards the positive or negative label. Over all samples this results in the shown density distinguishing higher and lower feature values by colour.

## Conclusion

We present a new approach to quantify dynamic one-year mortality risk in patients with MDS using gradient boosted decision tree ensembles. Based on two validation cohorts and cross validation on the training set we can show the advantages of using longitudinal features outperforming a purely diagnosis-based model with improvements among all evaluated scores. Longitudinal data adds important information to the prediction from early on. Additionally, we can show that the model does not only learn and perform on a specific dataset but does generalize to other, independent datasets. It achieves this by learning meaningful label-feature relations correlating to known medical factors and disease markers.

Even beyond the learned label the model does pick up on adverse events like AML progression and transfusion burden. Even with three university clinics participating the number of samples limits the interpretability of results and our results should be seen as a proof-of-concept. For a fully validated and clinically usable model, more data from more centres from more countries would be needed.

The model is applicable in different environments shown by the federated validation on the external datasets.

We hope to further validate and improve the approach looking at other clinical endpoints of interest like disease progression or therapy onset. Due to the dynamic and easily adjustable pipeline, other features both at diagnosis and longitudinal can be tested for predictive power. Inclusion of both machine learning and longitudinal features into the clinical routine to support decision-making and analyse large amounts of personalized data are crucial to improve patient care and survival.

## Data Availability

Data and models used in the present work are not available online due to privacy concerns. An interactive demo of the models and all code is available online.

https://dietrichlab.de/PythonApps/dynamic_mds_paper/

https://github.com/dietrichlab-cs/dynamic_one-year_mortality_mds

https://doi.org/10.5281/zenodo.16102058

## SUPPLEMENTARY MATERIAL

### Dataset characteristics

**Supplementary Table A:** Dataset characteristics for the different data sources. For Heidelberg and Mannheim no cut-off of the last 60 days is performed. The main objective of the validation datasets is to show performance on real-life data without lots of curation and cherry-picking. Still, we see similar median demographics for all datasets with differing label ratios especially for the Mannheim cohort. The Heidelberg cohort has an overall shorter average follow-up and higher mortality indicating more high-risk patients.

| **Characteristic** | **Düsseldorf** | **Heidelberg** | **Mannheim** |
| --- | --- | --- | --- |
| Patients | 1024 | 286 | 31 |
| Age (diagnosis), median | 69 | 72 | 72 |
| Follow-up years, median | 5 | 2 | 4.95 |
| Observation span | 1990-2022 | 2002-2025 | 2002-2015 |
| Gender |  |  |  |
| Female | 391 | 100 | 18 |
| Male | 633 | 186 | 13 |
| Mortality during follow-up | 658 (64.3%) | 167 (58.3%) | 12 (38.7%) |
| AML progression | 282 (27.5%) | 68 (23.8%) | 5 (16.1%) |
| Samples | 6145 | 1708 | 237 |
| Average samples per patient | 6 | 6 | 7.6 |
| Label ratio | 2.66:1 | 3:1 | 10:1 |
| Maximum quarter sampled | 32 | 32 | 32 |

### Longitudinal feature extraction

**Supplementary Table B:**
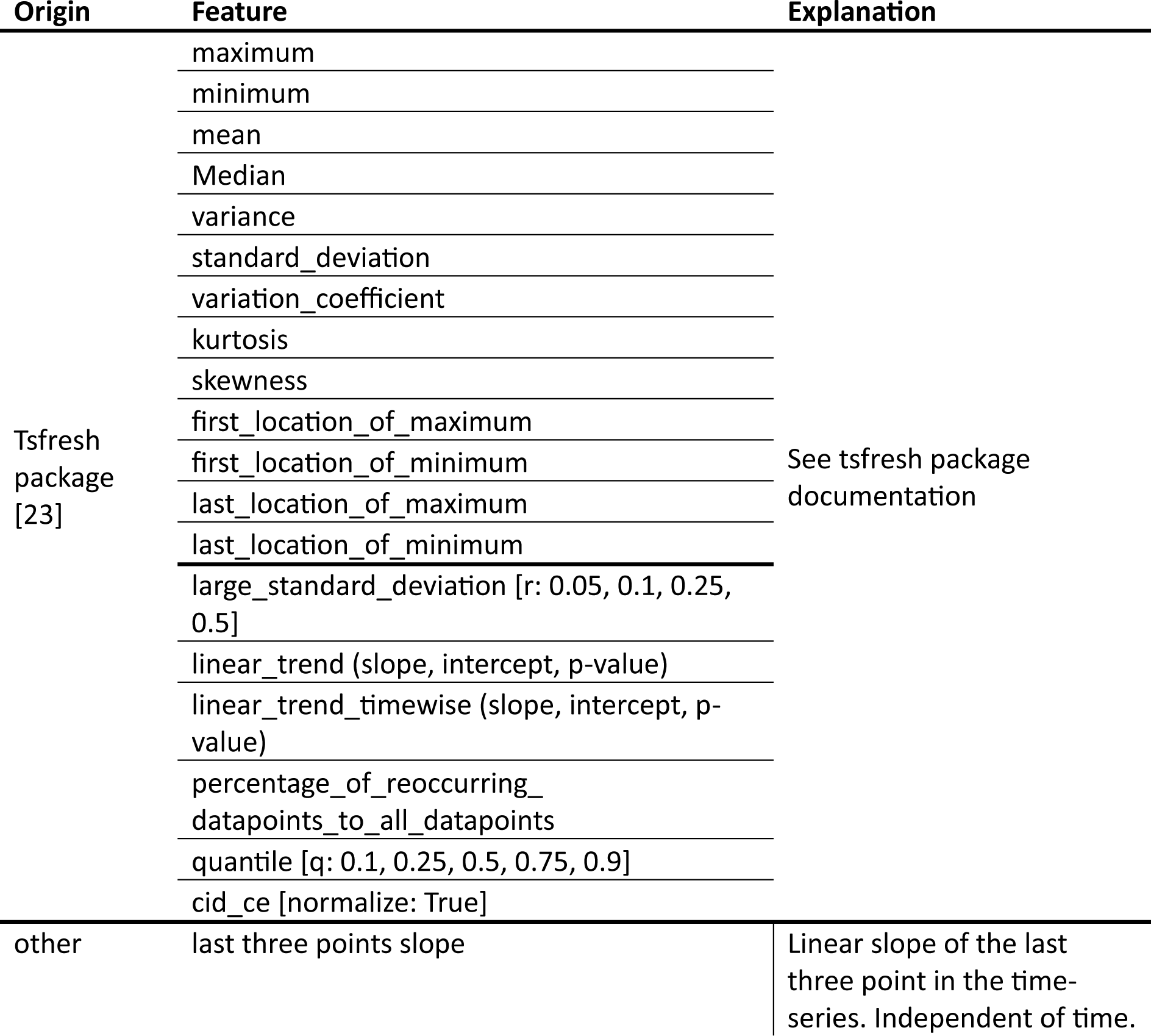
Features extracted from longitudinal data. Each feature is extracted for each sample and longitudinal parameter for the series of value from diagnosis to the point of prediction.

### Hyperparameters

**Supplementary Table C:** Hyperparameters for both models. Note that since we use XGBoost, features_per_split are converted to a percentage (features_per_split/total_number_of_features).

| Hyperparameter | Baseline model | Longitudinal model |
| --- | --- | --- |
| n_estimators | 500 | 1500 |
| learning_rate | 0.01 | 0.01 |
| max_depth | 3 | 3 |
| features_per_split | all | 80 |
| random_state | 123 | 123 |
| sample_weight | 2.3 | 2.3 |

### Samples per quarter

**Supplementary Figure A:**
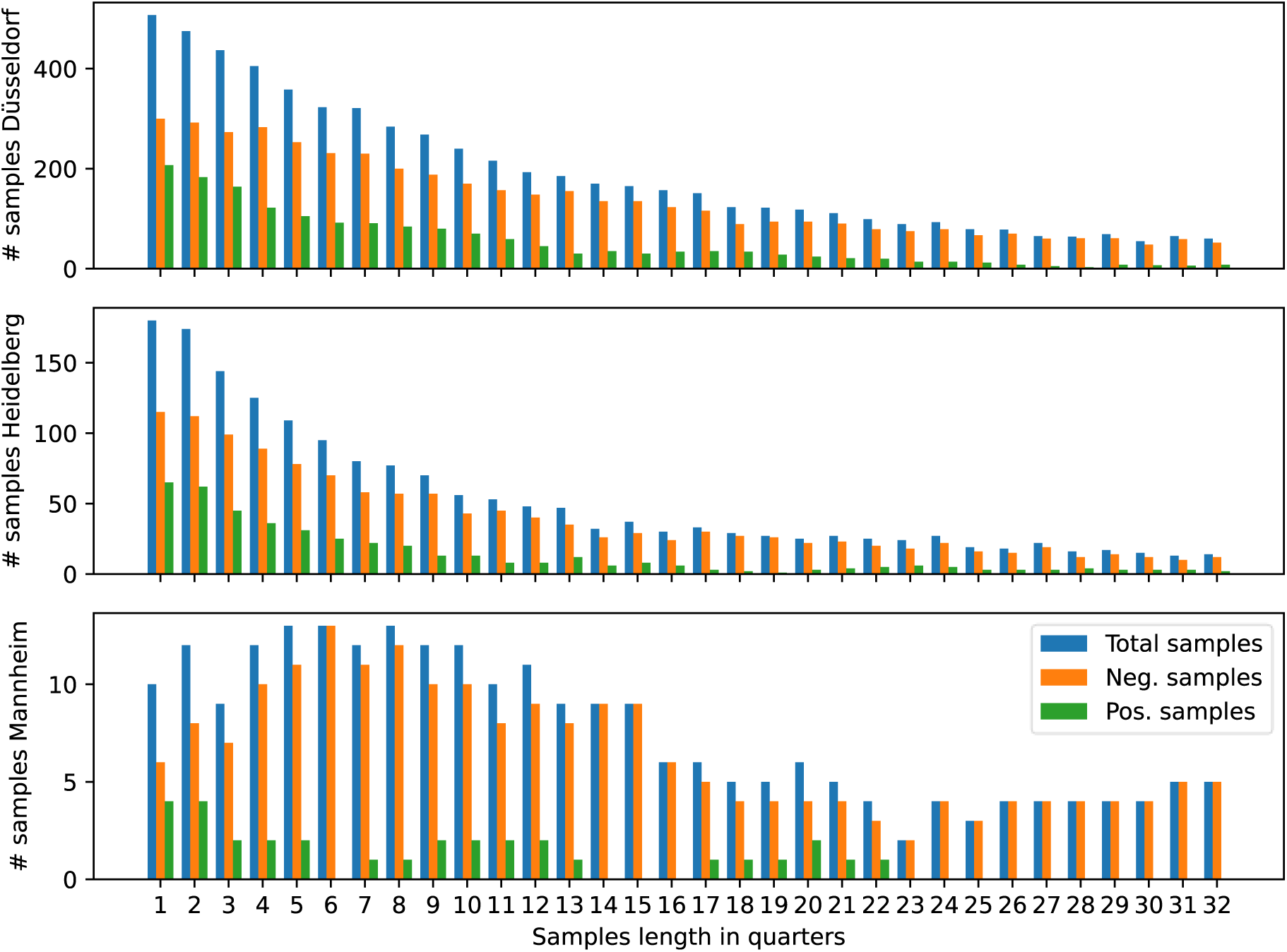
Sampling lengths over time given for each quarter. Negative samples have label 0 while positive ones have label 1.

**Supplementary Figure B:**
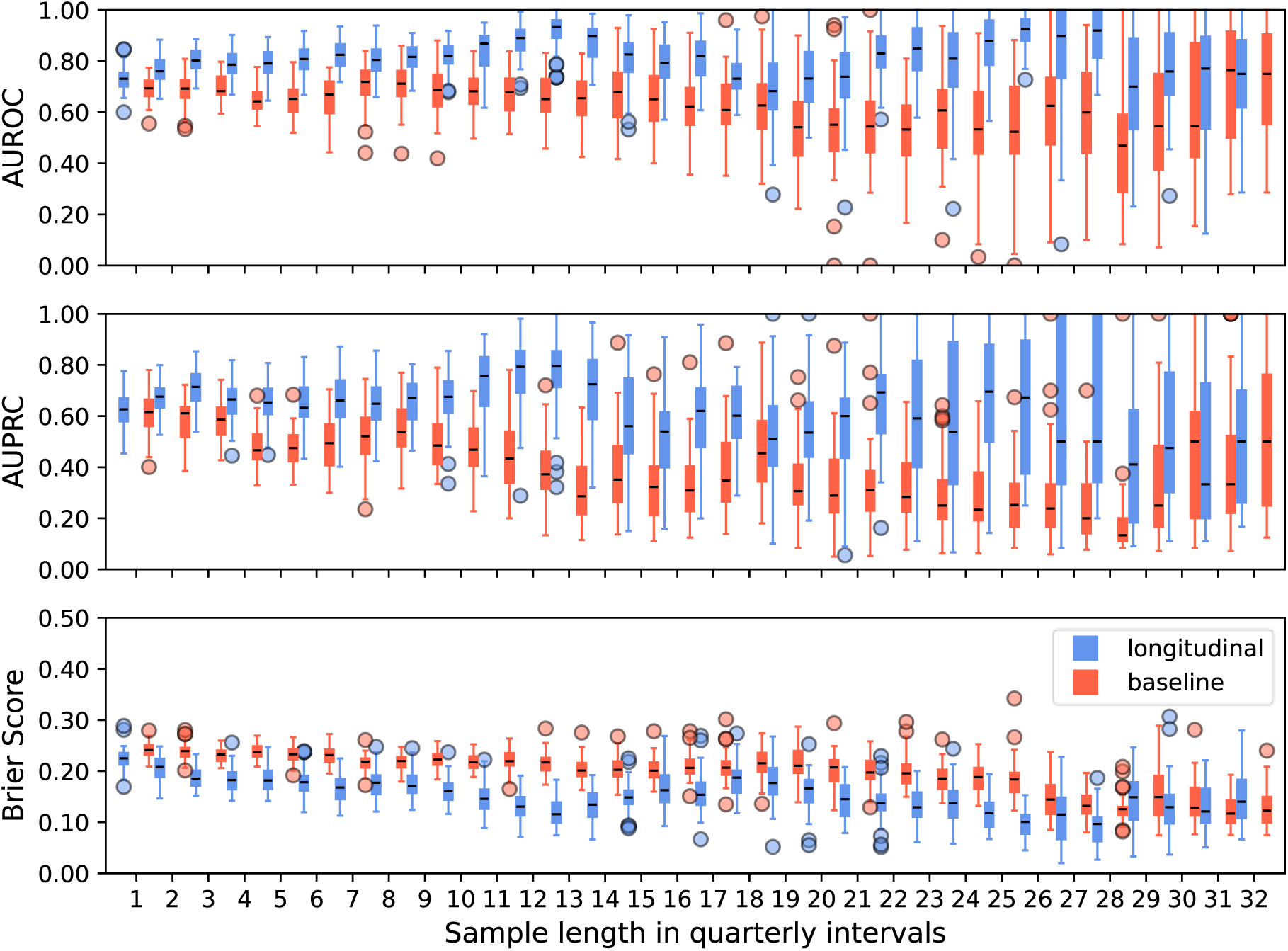
AUROC, AUPRC and Brier Score with variance bars on the cross-validation run on the Düsseldorf dataset. Bars represent inner quartile range (IQR) with the whiskers extending to 1.5xIQR. Outliers are denoted by circles.

**Supplementary Figure C:**
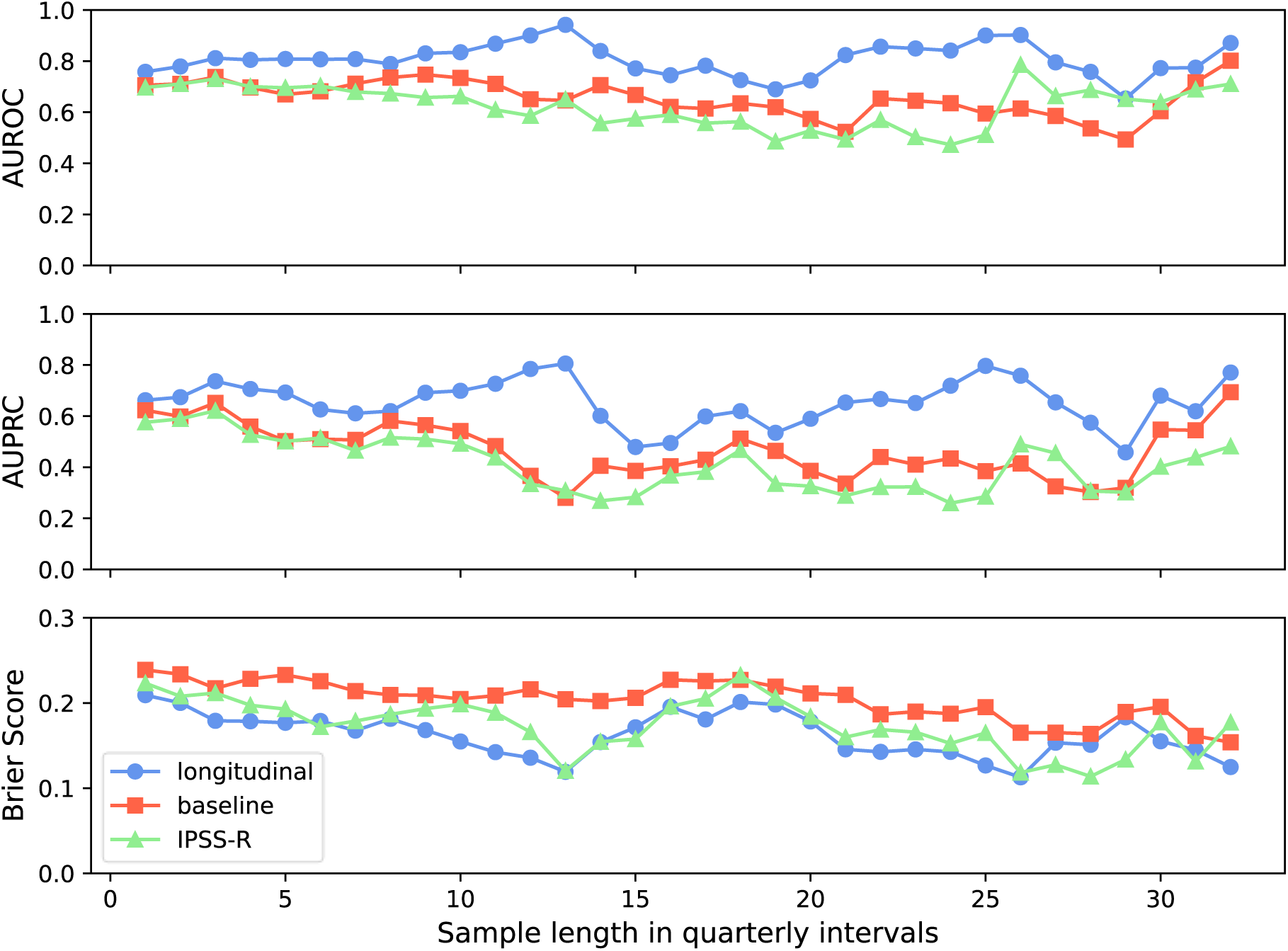
Comparison of average evaluation metrics over time between IPSS-R, using conditional Kaplan Meier estimators, and the baseline and longitudinal models. The dataset is based on all sample from the Düsseldorf dataset for patients with a present IPSS-R. This limits the dataset to n=689 patients.

## Acknowledgments

We would like to thank all members of the Algorithmic Bioinformatics and Hematology Research Lab groups at the Heinrich Heine University Düsseldorf for their feedback and discussions around the project. We also want to thank all clinical personnel and patients providing and processing data.

## Contributions

JB developed, evaluated and validated the software. UG, GK, PS and JB conceptualized the approach. PS, UG and GK supervised development and evaluation. SR and TL provided data for the Heidelberg validation cohort and supervised validation. SR ran the software in Heidelberg. AS, CG and NS provided data for the Mannheim validation cohort. AS and NS supervised validation. UG and CS provided data for the Düsseldorf cohort based on the MDS Registry Düsseldorf. AS, NS, TL, UG, CS, FS and SD provided detailed medical feedback. GK, PS and SR provided feedback on statistical analysis and model training. All authors have contributed to the internal review process, read and approved the manuscript.

